# Text Message intervention to improve function in people with low back pain (TEXT4myBACK): Statistical Analysis Plan

**DOI:** 10.1101/2025.06.03.25328920

**Authors:** Sana Shan, Laurent Billot, Manuela Ferreira

## Abstract

The TEXT4myBACK trial aims to examine the effect of lifestyle-focused program delivered by text message on function in people with non-persistent low back pain. The lifestyle-focused intervention is delivered as a mobile phone text message over a period of 12 weeks. The statistical analysis plan pre-specifies the method of analysis for every outcome and key variables collected in the trial.

The primary outcome is a composite endpoint incorporating measures of pain, functional status, patient reported impression of change, generic quality of life, levels of activity, hospital as well as medication use at 3-months follow-up that will be categorised into a hierarchy with number one ranked as most important according to clinical importance. The primary composite endpoint will be analysed using a hierarchical win ratio approach. Win-ratio approach maintains sequential assessment of outcomes while simultaneously assessing the effect of intervention for multiple outcomes as a single measure.

Secondary, exploratory and subgroup group analysis have been pre-specified as well.

## 1 Administrative information

### 1.1 Study identifiers

Australian New Zealand Clinical Trials Registry: ACTRN12618001263280, Date: 26 July 2018 NSLHD reference: RESP/19/005, ETH 13895

### 1.2 Revision history

| Version | Date | Details |
| --- | --- | --- |
| 0.1 (draft) | 10 March 2025 | First draft by Sana |
| 0.2 (draft) | 02 April 2025 | Second draft by Sana |
| 1.0 | 04 June 2025 | Final draft by Sana |

### 1.3 Contributors to the statistical analysis plan

SS – Main author; developed the initial draft and prepared subsequent versions. LB, MF - Reviewed every draft and revision.

## 2 Introduction

### 2.1 Study synopsis

The TEXT4myBACK trial^1^ is a two-arm, parallel, double blinded (assessor and statistician blinded) randomised controlled trial designed to evaluate the effectiveness of a self-management text message intervention compared to minimal intervention in improving function in adults living in the community with non-persistent low back pain (LBP). The lifestyle-focused intervention is delivered as a mobile phone text message over a period of 12 weeks. It includes content incorporating education and advice on healthy lifestyle behaviours, including physical activity, mood and sleep, along with medication and use of care. Evidence-based and disease-specific education will also be provided, including information on the benign nature of the condition, the limited role of imaging, complex interventions (i.e. surgery) and strong analgesics (e.g. opioids). The development of the intervention in collaboration with the TEXT ME^2^ research team, involvement of expert and consumer reviews during development phase as well as conducting a pilot study to test the feasibility of the identification and recruitment methods for both pharmacists and participants, have been discussed in detail in the protocol^1^.

### 2.2 Study population

People living in Australia, who will contact the research team with non-persistent LBP (i.e. less than 12 weeks duration) and meet the study’s inclusion criteria will be invited to participate. A total of 304 participants will be randomised from the community, with recruitment commencing in the Sydney Metropolitan area and expanding to NSW and other states in Australia.

#### 2.2.1 Inclusion Criteria

To be included, individuals must meet all the following inclusion criteria:

- Be aged 18 years or older.
- Have an episode of non-specific LBP of less than 12 weeks duration, with or without the presence of leg pain.
- Have pain classified at least as ‘moderate’ or above in the pain scale of the SF-12:

- During the past week, how much did pain interfere with your normal work (including work outside the home and housework)? 1-Not at all, 2-A little bit, 3-Moderately, 4-Quite a bit, 5-Extremely
- Familiarity with the use and access to, a phone that can receive text messages.

#### 2.2.1 Exclusion Criteria

Patients are not eligible if they meet any of the following criteria:

- Presence of serious pathology (described in detail in protocol^1^).
- Current pregnancy.
- Spinal surgery within the preceding 12 months.
- Co-morbid health conditions that would prevent active participation in the physical activity programs (e.g. unstable angina, uncontrolled hypertension).
- Inadequate English to understand the text messages or complete outcome measures.
- Any disorder/reason that may reduce capacity to understand and give informed consent.

## 3 Study interventions

All participants including those in the intervention and control groups will receive a welcome text informing that all study text messages will be signed as ‘#usyd’ and acknowledging their participation. All participants will receive text messages containing links to the online outcome measure surveys, utilisation of health care surveys and adverse events surveys at the specified time points.^1^ These messages will advise participants to contact the research team via the study email or phone number if they feel unsafe or uncertain about their LBP. At 12 weeks, all participants will receive a text message informing them that they will no longer receive text messages with information about low back pain, but they will still receive links to complete the study surveys.

In addition to the above-mentioned text messages, participants randomised to the control (minimal intervention) group will receive a text message (and an email if the participant does not use a smartphone) containing a link to access a LBP and healthy diet information package in REDCap.

### 3.1 Randomisation

Randomisation will be provided off-site using the REDCap software to the intervention or control group in a 1:1 ratio. The participants will be randomised after they signed the consent form and completed the baseline survey. The TEXT ME team health counsellor will perform the randomisation once per week and will be the only person other than the trial coordinator who will be aware of the group allocation of the participant. The trial coordinator will also be responsible for uploading the participant’s details into the text message software.

### 3.2 Study treatment

All consenting participants randomised to the intervention group will receive 4 text messages every week for the 12-week duration of the intervention period. These text messages will be semi-personalised providing advice, motivation and information to improve physical activity, sleep, mood, use of care and medication. Personalisation features include using participant’s preferred contact name, as well as sub-classification (e.g. messages from sleep domain will only be sent to those who reported sleep issue). Text message delivery will be managed by the TEXT ME software program and is designed to be delivered one-way (to the participant only). The software program will record all text message replies from participants for safety and potential descriptive analysis of their content. If the participant wishes to stop receiving text messages, they will be informed that they can reply “STOP’ at any time. Furthermore, participants in the intervention group will receive reminders (via text messages) every 4 weeks to complete the use of care surveys online. Detailed description is available in the protocol^1^ on the intervention including key domains, days and different time slots as well as delivery time of specific domain messages.

## 4 Outcomes

### 4.1 Primary outcome

The primary outcome is a composite outcome of the following variables at 3-months follow-up that will be categorised into a hierarchy (as shown below) with number one ranked as most important according to clinical importance:

1. Overall self-reported health score (visual analogue scale VAS) from EQ5D-5L questionnaire
2. Overall mobility measured on Likert scale from EQ-5D-5L questionnaire
3. Average pain intensity in the past week
4. Ability to perform daily activities from physical function scale 0-30
5. Amount of physical activity engaged within the past week (total time of moderate-vigorous physical activity in the previous week from Active Australia questionnaire)
6. Number of visits made to healthcare professionals in the past 12 weeks for low back pain
7. Any side or unpleasant effects experienced at 3 months (number of reported AEs)
8. Number of hours spent sitting or being inactive in a normal week (Average sum of time spent in 9 sedentary activities in a week in Sedentary Behaviour Questionnaire ((Typical weekday hours x 5) + (Typical weekend hours x 2) / 7)
9. The perceived overall change in symptoms from Global impression of change questionnaire
10. Amount of pain medicines taken in the past 12 weeks to manage low back pain
11. Anxiety and depression linked to low back pain measured on Likert scale from EQ-5D-5L

### 4.2 Secondary outcomes

The secondary outcomes will include following outcomes at 3, 6 and 12 months:

- Health state (utility score) calculated from EQ5D-5L using value sets for Australian population.
- Average pain intensity in the past week
- Ability to perform daily activities from physical function scale
- Amount of physical activity engaged with in the past week as a binary variable from Active Australia Insufficiently Active (1-149 min/week) and Sufficiently Active (≥150 min/week))
- The perceived overall change in symptoms from Global impression of change (<0 as worse, 0 as no change, >0 as improvement)
- Hours spent per week in sedentary behaviour from Sedentary behaviour questionnaire
- eHealth literacy questionnaire

### 4.3 Safety outcomes

- Smallest worthwhile effect stated as a baseline survey to analyse whether the score they achieve at 3 months is meaningful to the individual
- Health care utilisation survey asking about any referrals to health professionals or other services and medications prescribed or purchased related to their LBP every 4 weeks

### 4.4 Safety outcomes

- Occurrence of any adverse event (AE) or serious adverse event (SAE) can be reported in the study outcome surveys completed at 3, 6 and 12 months.

## 5 Analysis principles

### 5.1 Sample size

A sample size of 152 per group (total of 304 participants) will provide 90% power to detect an effect size of 0.4, assuming a standard deviation (SD) of 7 and allowing for a loss to follow-up rate of 15% at 3 months based on the between-group difference on our primary outcome patient specific functional scale (0-30 points) at 3 months (primary endpoint).

### 5.2 Software

Analyses will be conducted primarily using SAS Enterprise Guide (version 8.3 or above) and R (version 4.0.0 or above).

### 5.3 Interim analyses

No formal interim analyses were conducted during the study.

### 5.4 Multiplicity adjustment

Statistical tests are to be two-sided with a nominal level of α set at 5%. Analyses of the primary outcome (win ratio composite) will be unadjusted for multiplicity. Given that secondary outcomes are mostly components of the primary outcome, no adjustment for multiplicity will be applied. Instead, the analysis of secondary outcomes will be considered exploratory and used to describe the contribution of each component to the overall effect. Secondary outcomes will be analysed regardless of significance for the primary outcome.

### 5.5 Datasets analysed

#### 5.5.1 Analysis populations

The analysis population is intention-to-treat (ITT) population. The intention-to-treat (ITT) analysis set will be used to assess both effectiveness and safety.

#### 5.5.1 Analysis strategy

For all outcomes, the analyses will be performed on the ITT population using all available data. It is expected that the rate of missingness will be around 15%, multiple imputation techniques will be considered if missingness is > 15% as part of the sensitivity analysis to impute missing data.

Participants who withdraw or are lost to follow-up will not be replaced but their available data will be included in an intention to treat analysis unless specified prior to analysis. By withdrawing from the study, no further data will be collected, and no further contact will be made with the participants.

## 6 Planned Analyses

### 6.1 Subject disposition

The flow of patients through the trial will be displayed in a CONSORT^3^ (Consolidated Standards of Reporting Trials) diagram [see Figure 1]. The report will include the following: the number of participants assessed for eligibility, reasons for exclusion, the number of enrolled participants, the number available at 3, 6 and 12-months follow-up timepoints within the intervention and control arm. The flow chart will also include the number of participants lost to follow-up/ withdrew from the study as well as the reason for withdrawal at each timepoint.

### 6.2 Baseline comparison

#### 6.2.1 Patient characteristics

Description of baseline patient characteristics will be presented by treatment group. Discrete variables will be summarised by frequencies and percentages. Percentages will be calculated according to the number of patients in whom data are available. Continuous variables will be summarised by using mean and SD, and median and interquartile range (Q1-Q3). Baseline measures for all patients will be tabulated for the variables listed below

- Anthropometrics: age (years), sex, weight (kg), height (cm) and BMI (kg/m^2^)
- Demographics and socio-economic status including education, marital status, employment status
- Sleep issues, medication use
- LBP duration and symptom distribution
- Self-Administered Comorbidity Questionnaire – This is a validated instrument for the assessment of comorbidities^4^, which has been previously used to record comorbidities of patients with LBP^5, 6^

### 6.3 Follow-up assessments

All assessments performed and interventions received during follow-up will be described by treatment group. Details are presented in mock tables. No formal statistical tests are planned for these variables.

### 6.4 Analysis of the primary outcome

The primary composite endpoint will be analysed using a hierarchical win ratio approach. Win-ratio approach maintains sequential assessment of outcomes while simultaneously assessing the effect of intervention for multiple outcomes as a single measure.

#### 6.4.1 Main analysis

The outcomes will be ranked to reflect their clinical importance. A threshold will be defined indicating a clinically meaningful difference for each outcome. The decision process is explained in the table below. All participants in the intervention group will be paired with every participant in the control group (152 x 152 = 23,104 pairs). Within each pair, the response to each outcome will be compared in a hierarchical fashion to determine whether it is a win, a loss or a tie. If there’s no difference in response to a particular outcome within that pair, the decision is based on the response to the next ranked outcome.

Across all pairs, the ratio of wins and losses will be obtained. The results will be presented as win ratio (*R_W_*) with 95% confidence intervals (CI) obtained using Finkelstein and Schoenfeld^7^ method as explained in detail by Boentert et al^8^. “The total number of participants is represented by *N= N + N_C_*. For all possible pairs, the winner is assigned a score of 1, −1 or 0 based on a win, a loss or a tie respectively. The scores from all pairs are summed up for each participant with a positive integer *U_i_* indicating more wins than losses and vice versa. T statistic is calculated using a formula: 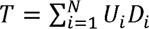 where Di = 1 if participant i is on the treatment and Di= 0 if participant i is on the control. Under the null hypothesis of no true difference between treatment and control, T has variance V where:

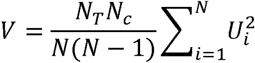

If T follows a normal distribution, z-score can be derived as *z*= *T*/√*V*, and obtain the p-value for a two-sided test for T given by 2xP(Z > z), where Z follows a standard normal distribution.

An approximate 95% CI for *R_W_* can also be calculated based on the z-score by first deriving the standard error (SE) for log *R_W_*:

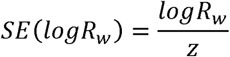

The 95% CI around 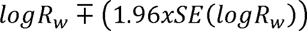. The limits are then exponentiated to derive CI around *R_W_*.”

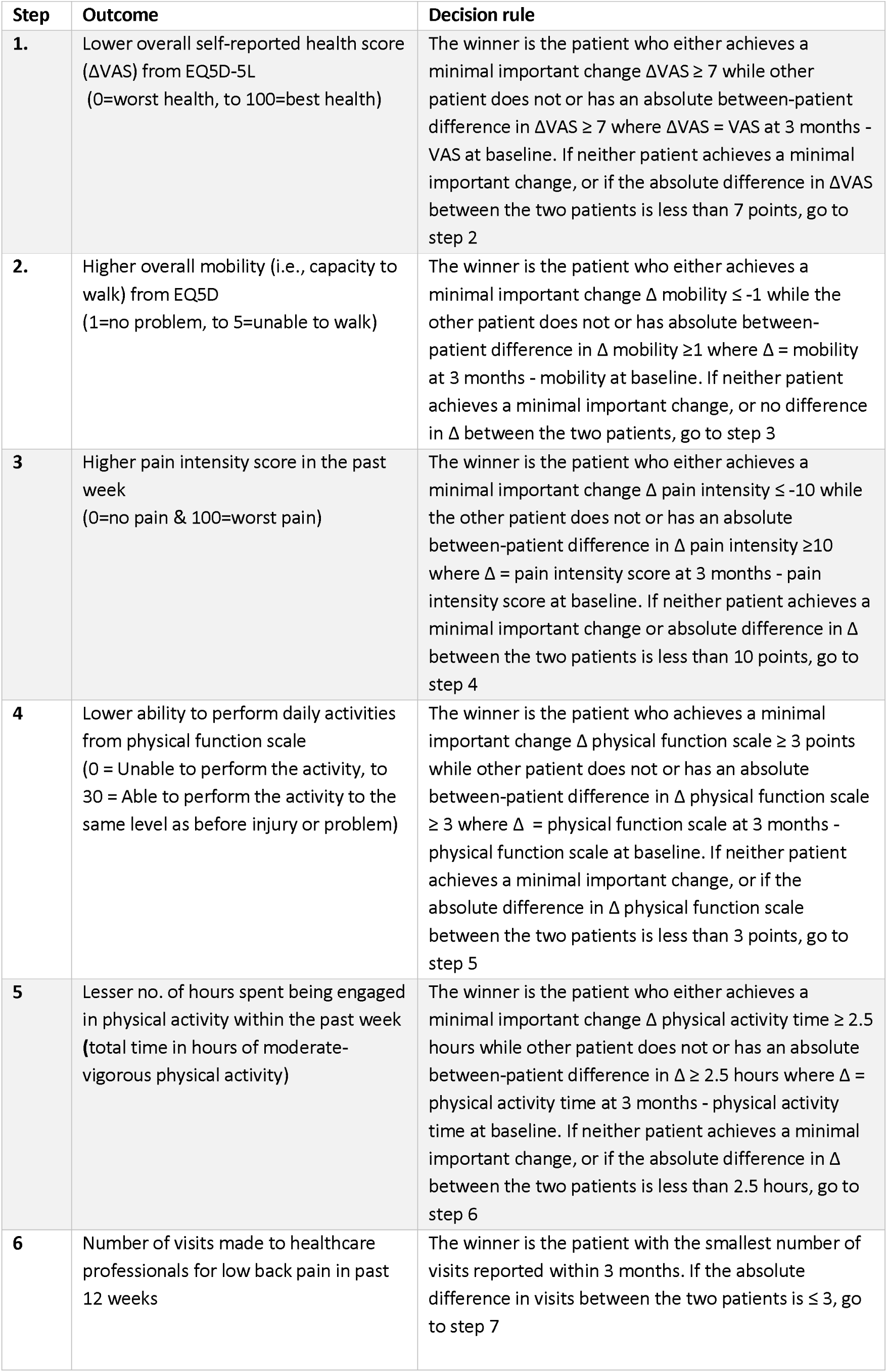
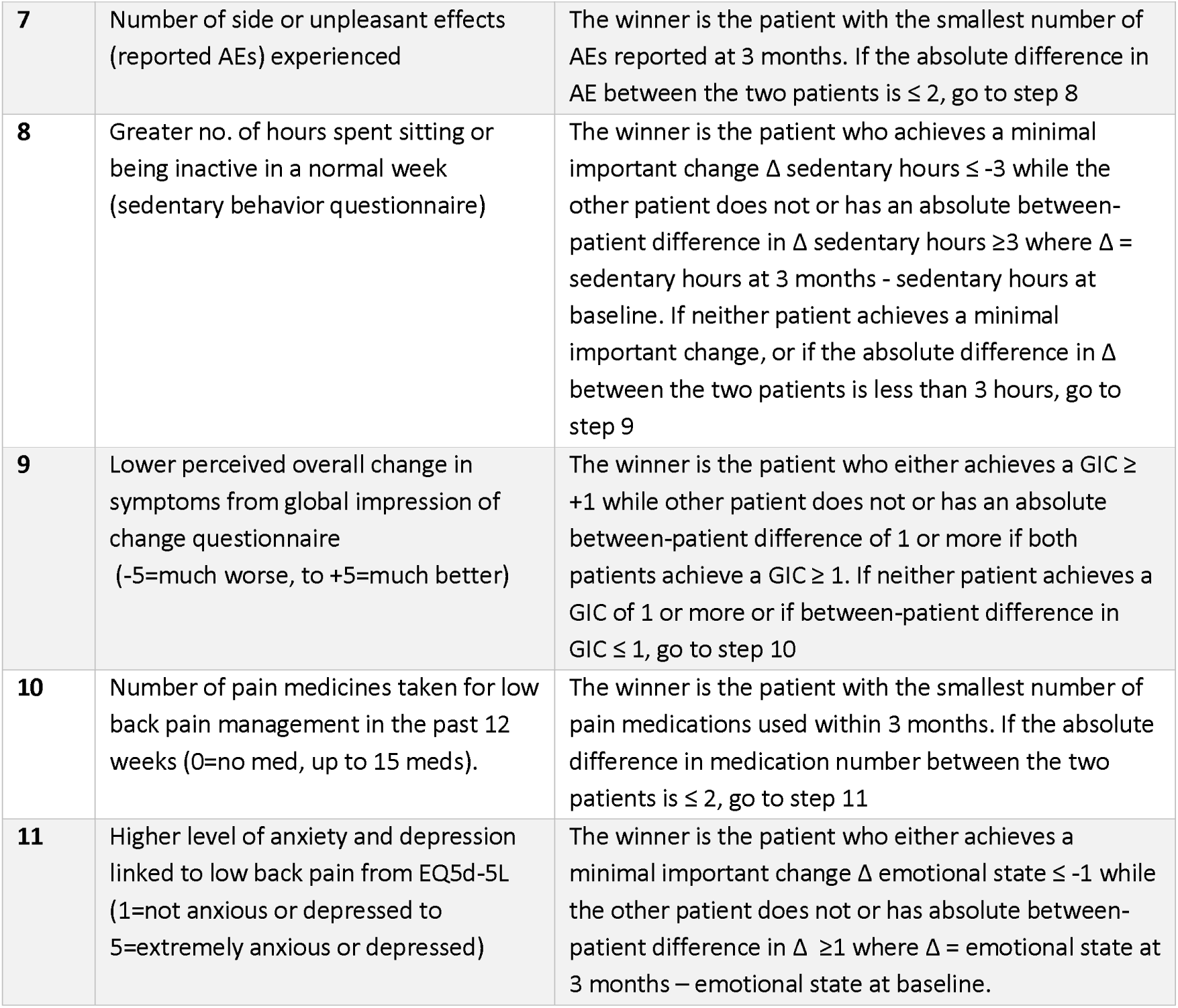

| Step | Outcome | Decision rule |
| --- | --- | --- |
| 1. | Lower overall self-reported health score (ΔVAS) from EQ5D-5L<br>(0=worst health, to 100=best health) | The winner is the patient who either achieves a minimal important change ΔVAS $\geq 7$ while other patient does not or has an absolute between-patient difference in ΔVAS $\geq 7$ where ΔVAS = VAS at 3 months - VAS at baseline. If neither patient achieves a minimal important change, or if the absolute difference in ΔVAS between the two patients is less than 7 points, go to step 2 |
| 2. | Higher overall mobility (i.e., capacity to walk) from EQ5D<br>(1=no problem, to 5=unable to walk) | The winner is the patient who either achieves a minimal important change $\Delta$ mobility $\leq -1$ while the other patient does not or has absolute between-patient difference in $\Delta$ mobility $\geq 1$ where $\Delta$ = mobility at 3 months - mobility at baseline. If neither patient achieves a minimal important change, or no difference in $\Delta$ between the two patients, go to step 3 |
| 3 | Higher pain intensity score in the past week<br>(0=no pain & 100=worst pain) | The winner is the patient who either achieves a minimal important change $\Delta$ pain intensity $\leq -10$ while the other patient does not or has an absolute between-patient difference in $\Delta$ pain intensity $\geq 10$ where $\Delta$ = pain intensity score at 3 months - pain intensity score at baseline. If neither patient achieves a minimal important change or absolute difference in $\Delta$ between the two patients is less than 10 points, go to step 4 |
| 4 | Lower ability to perform daily activities from physical function scale<br>(0 = Unable to perform the activity, to 30 = Able to perform the activity to the same level as before injury or problem) | The winner is the patient who achieves a minimal important change $\Delta$ physical function scale $\geq 3$ points while other patient does not or has an absolute between-patient difference in $\Delta$ physical function scale $\geq 3$ where $\Delta$ = physical function scale at 3 months - physical function scale at baseline. If neither patient achieves a minimal important change, or if the absolute difference in $\Delta$ physical function scale between the two patients is less than 3 points, go to step 5 |
| 5 | Lesser no. of hours spent being engaged in physical activity within the past week<br>(total time in hours of moderate-vigorous physical activity) | The winner is the patient who either achieves a minimal important change $\Delta$ physical activity time $\geq 2.5$ hours while other patient does not or has an absolute between-patient difference in $\Delta \geq 2.5$ hours where $\Delta$ = physical activity time at 3 months - physical activity time at baseline. If neither patient achieves a minimal important change, or if the absolute difference in $\Delta$ between the two patients is less than 2.5 hours, go to step 6 |
| 6 | Number of visits made to healthcare professionals for low back pain in past 12 weeks | The winner is the patient with the smallest number of visits reported within 3 months. If the absolute difference in visits between the two patients is $\leq 3$, go to step 7 |
| **7** | Number of side or unpleasant effects (reported AEs) experienced | The winner is the patient with the smallest number of AEs reported at 3 months. If the absolute difference in AE between the two patients is $\leq 2$, go to step 8 |
| **8** | Greater no. of hours spent sitting or being inactive in a normal week (sedentary behavior questionnaire) | The winner is the patient who achieves a minimal important change $\Delta$ sedentary hours $\leq -3$ while the other patient does not or has an absolute between-patient difference in $\Delta$ sedentary hours $\geq 3$ where $\Delta$ = sedentary hours at 3 months - sedentary hours at baseline. If neither patient achieves a minimal important change, or if the absolute difference in $\Delta$ between the two patients is less than 3 hours, go to step 9 |
| **9** | Lower perceived overall change in symptoms from global impression of change questionnaire (-5=much worse, to +5=much better) | The winner is the patient who either achieves a GIC $\geq +1$ while other patient does not or has an absolute between-patient difference of 1 or more if both patients achieve a GIC $\geq 1$. If neither patient achieves a GIC of 1 or more or if between-patient difference in GIC $\leq 1$, go to step 10 |
| **10** | Number of pain medicines taken for low back pain management in the past 12 weeks (0=no med, up to 15 meds). | The winner is the patient with the smallest number of pain medications used within 3 months. If the absolute difference in medication number between the two patients is $\leq 2$, go to step 11 |
| **11** | Higher level of anxiety and depression linked to low back pain from EQ5d-5L (1=not anxious or depressed to 5=extremely anxious or depressed) | The winner is the patient who either achieves a minimal important change $\Delta$ emotional state $\leq -1$ while the other patient does not or has absolute between-patient difference in $\Delta \geq 1$ where $\Delta$ = emotional state at 3 months – emotional state at baseline. |

### 6.5 Analysis of the secondary outcomes

For continuous outcomes including patient-specific functional scale, average pain intensity during the past week, Health utility score from EQ5D and hours per week spent in sedentary behaviour, linear mixed models will be used to assess the effect of the treatment group. The model will include outcome data collected at every post-baseline visit, i.e. at months 3, 6 and 12. Fixed effects will include the randomised treatment allocation, the follow-up time points as a categorical variable with 3 levels, the interaction between treatment and time-point as well as the baseline measurement of the outcome. Within-patient correlations will be modelled using a repeated patient effect. The overall effect of the intervention will be estimated as the mean difference and 95% CI between the intervention and control arm over the entire follow-up i.e. by combining data obtained across all visits. The effect of the intervention at specific time points (e.g. at Month 12) will be estimated from the same model using contrasts.

The outcomes of participant’s global impression of change (<0 as worse, 0 as no change, >0 as improvement) will be analysed on an ordinal scale. The distribution of ordinal outcome variable between randomised groups will be analysed at 3, 6 and 12 months using a mixed model with repeated measures (as mentioned above) with multinomial distribution and a cumulative logit link function. The overall effect of the intervention will be estimated as odds ratio with 95% CI. Similar approach will be used for the analysis of binary variable of physical activity engagement (sufficiently active vs. insufficiently active) with binomial distribution and logit link function.

The distribution of eHealth literacy outcome related to knowledge and confidence in finding and understanding digital health information will be analysed descriptively for each follow-up timepoint for 7 independent scales from eHLQ questionnaire with averaged score ranging from 1 – 4. Each scale consists of 4 – 6 items with assigned value 1 strongly disagree – 4 strongly agree. The results will be presented as mean and standard deviation.

### 6.6 Sensitivity analyses

No Sensitivity analyses are planned.

### 6.7 Treatment of missing data

To evaluate the impact of missing data, multiple imputation^9^ will employed generating five datasets to estimate changes from baseline at 3 months for all randomised participants. The imputation model will include age at baseline, sex, treatment group, baseline values of outcome measures (listed in section 5.1) as well their follow-up values at 3, 6 and 12 months. Win ratios will be calculated separately for each imputed dataset using the same approach a mentioned in section 7.4.1, the results will be pooled to produce a single overall estimate.

### 6.8 Per-protocol analysis

No per-protocol analysis will be carried out.

### 6.9 Analysis of the exploratory outcomes

Analysis of exploratory outcomes including smallest worthwhile effect (SWE) (binary outcome of whether smallest worthwhile effect achieved or not) at month 3 and health utilisation over 12 months will be analysed using descriptive statistics. Using a heatmap, the trends in healthcare utilisation (counts of visits to healthcare professionals) will be visualised for each participant over time (at every 4 weeks from baseline to 12 months). Analysis of MBS and PBS data will be analysed at a later stage and will not be included as part of this analysis.

### 6.10 Analysis of safety outcomes

Safety outcomes of all-cause and cause-specific SAEs will be summarised as the number and proportion of patients experiencing at least one event. Adverse events will be summarised by category of event and overall numbers of events. In addition to the number of patients with at least one event, we will report the total number of events.

A listing of all AEs and SAEs will be reported (in an appendix).

### 6.11 Subgroup analyses

Pre-specified subgroup analyses will be carried out, irrespective of whether there is a significant treatment effect on the primary outcome. These subgroup analyses will only be performed in the ITT population. Subgroups based on pre-randomisation patient characteristics assessed before randomisation are defined as follows:

- Age category (<65 vs. ≥65 years)
- Sex
- Baseline physical function score (<12 vs >=12)

Analysis for each subgroup will be performed by repeating the main analysis (as described in section 7.4.1) separately within each subgroup. The results will be displayed on a forest plot. No p-value for heterogeneity will be computed given that the analysis is only performed separately within each subgroup.

## Data Availability

The manuscript is a statistical analysis plan and does not contain any results.

## 8 Appendix 1: Proposed Tables and figures

**Figure 1:**
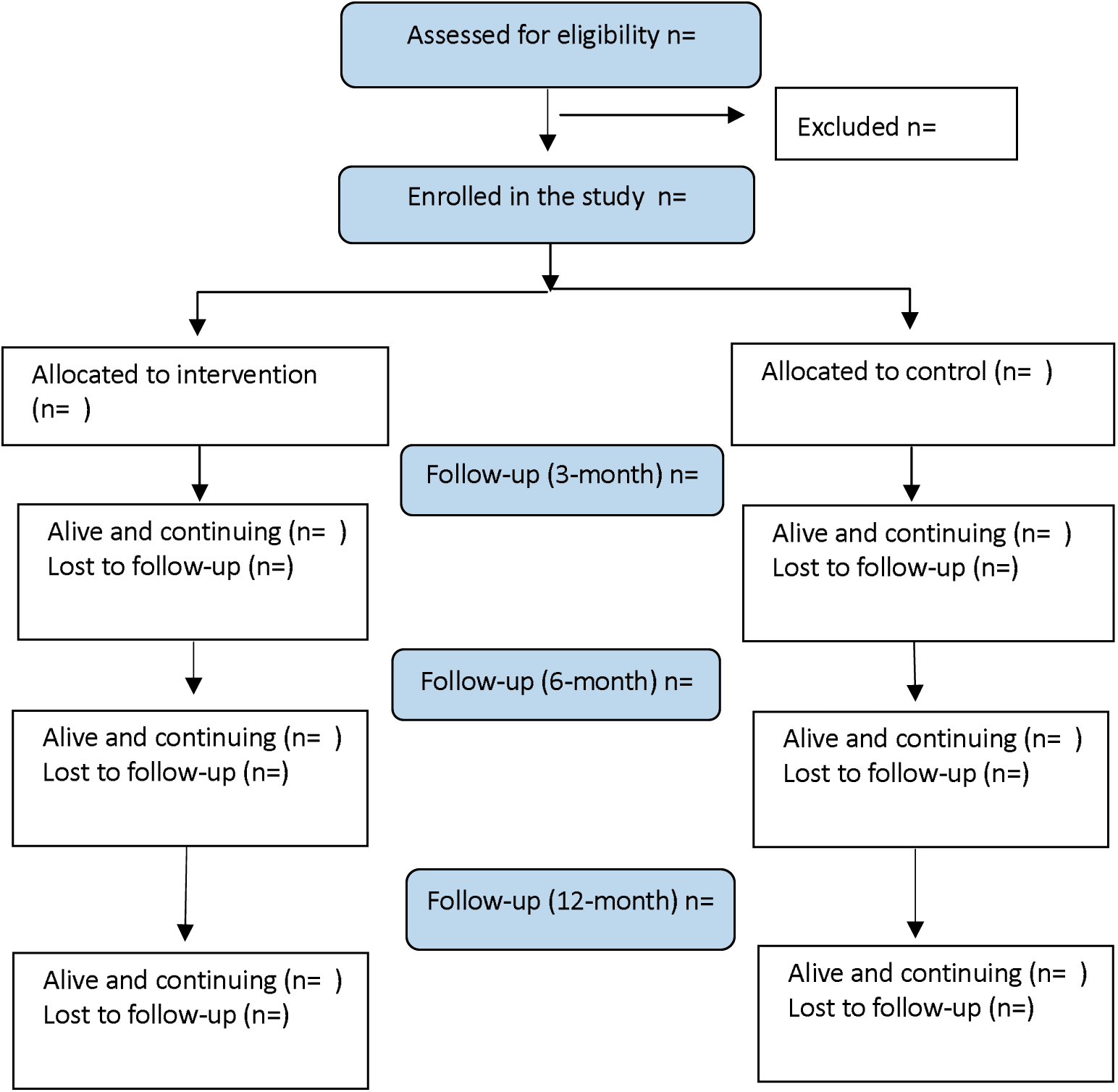
Consort flowchart

**Table 1.** Baseline characteristics.

|  | Intervention<br>(N = ) | Control<br>(N = ) | Total<br>(N = ) |
| --- | --- | --- | --- |
| **Age (years)** | xxx | xxx | xxx |
| Mean (SD) | xxx.x (xxx.x) | xxx.x (xxx.x) | xxx.x (xxx.x) |
| Median (Q1; Q3) | xxx (xxx; xxx) | xxx (xxx; xxx) | xxx (xxx; xxx) |
| min max | xxx to xxx | xxx to xxx | xxx to xxx |
| **Sex** | xxx | xxx | xxx |
| Male | xxx xx.x% | xxx xx.x% | xxx xx.x% |
| Female | xxx xx.x% | xxx xx.x% | xxx xx.x% |
| **Nationality** | xxx | xxx | xxx |
| xx | xxx xx.x% | xxx xx.x% | xxx xx.x% |
| xx | xxx xx.x% | xxx xx.x% | xxx xx.x% |
| **Living arrangement** |  |  |  |
| Single | xxx xx.x% | xxx xx.x% | xxx xx.x% |
| Married | xxx xx.x% | xxx xx.x% | xxx xx.x% |
| De-facto | xxx xx.x% | xxx xx.x% | xxx xx.x% |
| **Employment status** |  |  |  |
| Unemployed | xxx xx.x% | xxx xx.x% | xxx xx.x% |
| Part time | xxx xx.x% | xxx xx.x% | xxx xx.x% |
| Full time | xxx xx.x% | xxx xx.x% | xxx xx.x% |
| Volunteer | xxx xx.x% | xxx xx.x% | xxx xx.x% |
| Studying | xxx xx.x% | xxx xx.x% | xxx xx.x% |
| Full time carer (including parenting) | xxx xx.x% | xxx xx.x% | xxx xx.x% |
| Retired | xxx xx.x% | xxx xx.x% | xxx xx.x% |
| Other | xxx xx.x% | xxx xx.x% | xxx xx.x% |
| **Weight (kg)** | xxx | xxx | xxx |
| Mean (SD) | xxx.x (xxx.x) | xxx.x (xxx.x) | xxx.x (xxx.x) |
| Median (Q1; Q3) | xxx (xxx; xxx) | xxx (xxx; xxx) | xxx (xxx; xxx) |
| min max | xxx to xxx | xxx to xxx | xxx to xxx |
| **Height (cm)** | xxx | xxx | xxx |
| Mean (SD) | xxx.x (xxx.x) | xxx.x (xxx.x) | xxx.x (xxx.x) |
| Median (Q1; Q3) | xxx (xxx; xxx) | xxx (xxx; xxx) | xxx (xxx; xxx) |
| min max | xxx to xxx | xxx to xxx | xxx to xxx |
| **BMI** | xxx | xxx | xxx |
| Mean (SD) | xxx.x (xxx.x) | xxx.x (xxx.x) | xxx.x (xxx.x) |
| Median (Q1; Q3) | xxx (xxx; xxx) | xxx (xxx; xxx) | xxx (xxx; xxx) |
| min max | xxx to xxx | xxx to xxx | xxx to xxx |
| **Education level** | xxx | xxx | xxx |
| Elementary | xxx xx.x% | xxx xx.x% | xxx xx.x% |
| High School | xxx xx.x% | xxx xx.x% | xxx xx.x% |
| Graduate (TAFE, college) | xxx xx.x% | xxx xx.x% | xxx xx.x% |
| Graduate (Bachelor, Master) | xxx xx.x% | xxx xx.x% | xxx xx.x% |
| Doctorate (PhD) | xxx xx.x% | xxx xx.x% | xxx xx.x% |
| **Any job involving sitting or standing still for most of the work day** | xxx xx.x% | xxx xx.x% | xxx xx.x% |
| **Medical history** | xxx | xxx | xxx |
| Heart disease | xxx xx.x% | xxx xx.x% | xxx xx.x% |
| High blood pressure | xxx xx.x% | xxx xx.x% | xxx xx.x% |
| Lung disease | xxx xx.x% | xxx xx.x% | xxx xx.x% |
| Diabetes | xxx xx.x% | xxx xx.x% | xxx xx.x% |
| Ulcer or stomach disease | xxx xx.x% | xxx xx.x% | xxx xx.x% |
| Kidney disease | xxx xx.x% | xxx xx.x% | xxx xx.x% |
| Liver disease | xxx xx.x% | xxx xx.x% | xxx xx.x% |
| Anaemia or other blood disease | xxx xx.x% | xxx xx.x% | xxx xx.x% |
| Cancer | xxx xx.x% | xxx xx.x% | xxx xx.x% |
| Depression | xxx xx.x% | xxx xx.x% | xxx xx.x% |
| Osteoarthritis or degenerative arthritis | xxx xx.x% | xxx xx.x% | xxx xx.x% |
| Rheumatoid arthritis | xxx xx.x% | xxx xx.x% | xxx xx.x% |
| Other medical problems | xxx xx.x% | xxx xx.x% | xxx xx.x% |
| Any other injuries, conditions or diagnoses (e.g. knee pain, shoulder injury, heart disease, etc.) | xxx xx.x% | xxx xx.x% | xxx xx.x% |
| Sleep issues (i.e. difficulty getting to sleep or waking up at night) | xxx xx.x% | xxx xx.x% | xxx xx.x% |
| **Impact of COVID-19 on:** |  |  |  |
| Overall physical health |  |  |  |
| not at all | xxx xx.x% | xxx xx.x% | xxx xx.x% |
| a little | xxx xx.x% | xxx xx.x% | xxx xx.x% |
| moderate | xxx xx.x% | xxx xx.x% | xxx xx.x% |
| very much | xxx xx.x% | xxx xx.x% | xxx xx.x% |
| an extreme amount | xxx xx.x% | xxx xx.x% | xxx xx.x% |
| Overall mental health |  |  |  |
| not at all | xxx xx.x% | xxx xx.x% | xxx xx.x% |
| a little | xxx xx.x% | xxx xx.x% | xxx xx.x% |
| moderate | xxx xx.x% | xxx xx.x% | xxx xx.x% |
| very much | xxx xx.x% | xxx xx.x% | xxx xx.x% |
| an extreme amount | xxx xx.x% | xxx xx.x% | xxx xx.x% |
| Overall quality of life |  |  |  |
| not at all | xxx xx.x% | xxx xx.x% | xxx xx.x% |
| a little | xxx xx.x% | xxx xx.x% | xxx xx.x% |
| moderate | xxx xx.x% | xxx xx.x% | xxx xx.x% |
| very much | xxx xx.x% | xxx xx.x% | xxx xx.x% |
| an extreme amount | xxx xx.x% | xxx xx.x% | xxx xx.x% |

**Table 2.** Back pain assessment at baseline.

|  | Intervention<br>(N = ) | Control<br>(N = ) | Total<br>(N = ) |
| --- | --- | --- | --- |
| **Length of back pain symptoms** |  |  |  |
| Less than 6 weeks | xxx xx.x% | xxx xx.x% | xxx xx.x% |
| Greater than 6 weeks | xxx xx.x% | xxx xx.x% | xxx xx.x% |
| **First episode of low back pain** | xxx xx.x% | xxx xx.x% | xxx xx.x% |
| **Number of episodes of low back pain previously** | xx.x(xx.x) | xx.x(xx.x) | xx.x(xx.x) |
| **Pain extending to the buttock or leg that is related to your back pain?** | xxx xx.x% | xxx xx.x% | xxx xx.x% |
| **Pain in one or both legs** |  | xxx xx.x% | xxx xx.x% |
| One leg | xxx xx.x% | xxx xx.x% | xxx xx.x% |
| Both legs | xxx xx.x% | xxx xx.x% | xxx xx.x% |
| **Feeling tingling, pins and needles, or numbness in the leg(s)** | xxx xx.x% | xxx xx.x% | xxx xx.x% |
| **Presence of pain, numbness and/or** | xxx xx.x% | xxx xx.x% | xxx xx.x% |
| **fatigue in the legs that is worse in standing or walking and alleviated during flexion/sitting** |  |  |  |
| **Feeling loss of muscle strength in the legs** | xxx xx.x% | xxx xx.x% | xxx xx.x% |
| **Feeling difficulty in walking due to the pain in the leg(s) or back** | xxx xx.x% | xxx xx.x% | xxx xx.x% |
| **Cause of back pain (self-reported)** |  |  |  |
| An injury | xxx xx.x% | xxx xx.x% | xxx xx.x% |
| Accident | xxx xx.x% | xxx xx.x% | xxx xx.x% |
| Work related | xxx xx.x% | xxx xx.x% | xxx xx.x% |
| Congenital disorder | xxx xx.x% | xxx xx.x% | xxx xx.x% |
| No apparent cause | xxx xx.x% | xxx xx.x% | xxx xx.x% |
| Other | xxx xx.x% | xxx xx.x% | xxx xx.x% |
| **Sought professional help for back pain in the past 4 weeks** | xxx xx.x% | xxx xx.x% | xxx xx.x% |
| Acupuncture | xxx xx.x% | xxx xx.x% | xxx xx.x% |
| Chiropractor | xxx xx.x% | xxx xx.x% | xxx xx.x% |
| Exercise physiologist | xxx xx.x% | xxx xx.x% | xxx xx.x% |
| Massage therapist | xxx xx.x% | xxx xx.x% | xxx xx.x% |
| Pharmacist | xxx xx.x% | xxx xx.x% | xxx xx.x% |
| Orthotist | xxx xx.x% | xxx xx.x% | xxx xx.x% |
| Osteopath | xxx xx.x% | xxx xx.x% | xxx xx.x% |
| Physiotherapist | xxx xx.x% | xxx xx.x% | xxx xx.x% |
| General Practitioner (GP) | xxx xx.x% | xxx xx.x% | xxx xx.x% |
| Surgeon | xxx xx.x% | xxx xx.x% | xxx xx.x% |
| Medical specialist | xxx xx.x% | xxx xx.x% | xxx xx.x% |
| Other | xxx xx.x% | xxx xx.x% | xxx xx.x% |
| **Ave. out of pocket costs for these visits (AUD)** | xxx.x (xxx.x) | xxx.x (xxx.x) | xxx.x (xxx.x) |
| **Medicines purchased for back pain in the past 4 weeks (including all prescriptions, over the counter, alternative remedies, vitamins or supplements)(AUD)** | xxx.x (xxx.x) | xxx.x (xxx.x) | xxx.x (xxx.x) |
| **Hospital admissions or Emergency Department visits (back pain related) in the past 4 weeks** | xxx xx.x% | xxx xx.x% | xxx xx.x% |
| Number of days in hospital | xxx.x (xxx.x) | xxx.x (xxx.x) | xxx.x (xxx.x) |
| Attended a public or private hospital |  |  |  |
| Public | xxx xx.x% | xxx xx.x% | xxx xx.x% |
| Private | xxx xx.x% | xxx xx.x% | xxx xx.x% |
| Total of pocket costs for these visits? (i.e. how much was not covered by Medicare or Private health insurance in dollars? (AUD) | xxx.x (xxx.x) | xxx.x (xxx.x) | xxx.x (xxx.x) |
| Ave. out of pocket costs for these visits (AUD) | xxx.x (xxx.x) | xxx.x (xxx.x) | xxx.x (xxx.x) |

**Table 3.** Baseline assessment of all outcomes.

| Variable | Intervention<br>(N = ) | Control<br>(N = ) | Total<br>(N = ) |
| --- | --- | --- | --- |
| **Physical function, assessed using the Patient Specific Functional Scale (0-30)** | xxx | xxx | xxx |
| Mean (SD) | xxx.x (xxx.x) | xxx.x (xxx.x) | xxx.x (xxx.x) |
| Median (Q1; Q3) | xxx (xxx; xxx) | xxx (xxx; xxx) | xxx (xxx; xxx) |
| min max | xxx to xxx | xxx to xxx | xxx to xxx |
| **Low back pain intensity over the past week (0-100)** | xxx | xxx | xxx |
| Mean (SD) | xxx.x (xxx.x) | xxx.x (xxx.x) | xxx.x (xxx.x) |
| Median (Q1; Q3) | xxx (xxx; xxx) | xxx (xxx; xxx) | xxx (xxx; xxx) |
| min max | xxx to xxx | xxx to xxx | xxx to xxx |
| **Total time of moderate-vigorous physical activity in the previous week from Active Australia questionnaire (hours)** | xxx | xxx | xxx |
| Mean (SD) | xxx.x (xxx.x) | xxx.x (xxx.x) | xxx.x (xxx.x) |
| Median (Q1; Q3) | xxx (xxx; xxx) | xxx (xxx; xxx) | xxx (xxx; xxx) |
| min max | xxx to xxx | xxx to xxx | xxx to xxx |
| **Physical activity engagement from Active Australia questionnaire (ordinal classification)** |  |  |  |
| Insufficiently Active (1-149 min/week) | xxx xx.x% | xxx xx.x% | xxx xx.x% |
| Sufficiently Active (≥150 min/week) | xxx xx.x% | xxx xx.x% | xxx xx.x% |
| **Total time spent sitting or being inactive in a normal week from Sedentary behaviour questionnaire (hours)** | xxx | xxx | xxx |
| Mean (SD) | xxx.x (xxx.x) | xxx.x (xxx.x) | xxx.x (xxx.x) |
| Median (Q1; Q3) | xxx (xxx; xxx) | xxx (xxx; xxx) | xxx (xxx; xxx) |
| min max | xxx to xxx | xxx to xxx | xxx to xxx |
| **Participant's global impression of change (-5 to +5)** |  |  |  |
| Mean (SD) | xxx.x (xxx.x) | xxx.x (xxx.x) | xxx.x (xxx.x) |
| Median (Q1; Q3) | xxx (xxx; xxx) | xxx (xxx; xxx) | xxx (xxx; xxx) |
| min max | xxx to xxx | xxx to xxx | xxx to xxx |
| **Participant's global impression of change (-5 to +5)** |  |  |  |
| Worse (<= -1) | xxx xx.x% | xxx xx.x% | xxx xx.x% |
| No change (0) | xxx xx.x% | xxx xx.x% | xxx xx.x% |
| Improvement (>=+1) | xxx xx.x% | xxx xx.x% | xxx xx.x% |
| **Overall mobility from EQ5D-5L (1 to 5)** |  |  |  |
| Mean (SD) | xxx.x (xxx.x) | xxx.x (xxx.x) | xxx.x (xxx.x) |
| Median (Q1; Q3) | xxx (xxx; xxx) | xxx (xxx; xxx) | xxx (xxx; xxx) |
| min max | xxx to xxx | xxx to xxx | xxx to xxx |
| **Overall emotional health (Anxiety/Depression) from EQ5D-5L (1 to 5)** |  |  |  |
| Mean (SD) | xxx.x (xxx.x) | xxx.x (xxx.x) | xxx.x (xxx.x) |
| Median (Q1; Q3) | xxx (xxx; xxx) | xxx (xxx; xxx) | xxx (xxx; xxx) |
| min max | xxx to xxx | xxx to xxx | xxx to xxx |
| **Overall self-reported health score from EQ5D-5L (0 – 100)** |  |  |  |
| Mean (SD) | xxx.x (xxx.x) | xxx.x (xxx.x) | xxx.x (xxx.x) |
| Median (Q1; Q3) | xxx (xxx; xxx) | xxx (xxx; xxx) | xxx (xxx; xxx) |
| min max | xxx to xxx | xxx to xxx | xxx to xxx |
| **Health utility score from EQ5D-5L calculated for Australian value set** |  |  |  |
| Mean (SD) | xxx.x (xxx.x) | xxx.x (xxx.x) | xxx.x (xxx.x) |
| Median (Q1; Q3) | xxx (xxx; xxx) | xxx (xxx; xxx) | xxx (xxx; xxx) |
| min max | xxx to xxx | xxx to xxx | xxx to xxx |
| **Number of visits made in the past 4 weeks to healthcare professionals your low back pain** |  |  |  |
| Mean (SD) | xxx.x (xxx.x) | xxx.x (xxx.x) | xxx.x (xxx.x) |
| Median (Q1; Q3) | xxx (xxx; xxx) | xxx (xxx; xxx) | xxx (xxx; xxx) |
| min max | xxx to xxx | xxx to xxx | xxx to xxx |

| **Number of pain medicines taken for low back pain management in past 4 weeks** |  |  |  |
| --- | --- | --- | --- |
| Mean (SD) | xxx.x (xxx.x) | xxx.x (xxx.x) | xxx.x (xxx.x) |
| Median (Q1; Q3) | xxx (xxx; xxx) | xxx (xxx; xxx) | xxx (xxx; xxx) |
| min max | xxx to xxx | xxx to xxx | xxx to xxx |

**Table 4.** Descriptive statistics for eHealth Literacy Questionnaire at all time-points.

| **Outcome** | **Intervention**<br>**N Mean(SD)** | **Control**<br>**N Mean(SD)** |
| --- | --- | --- |
| **Scale** |  |  |
| **Baseline** |  |  |
| Using technology to process health information | XX xx.xx (xx.x) | XX xx.xx (xx.x) |
| Understanding of health concepts and language | XX xx.xx (xx.x) | XX xx.xx (xx.x) |
| Ability to actively engage with digital services | XX xx.xx (xx.x) | XX xx.xx (xx.x) |
| Feel safe and in control | XX xx.xx (xx.x) | XX xx.xx (xx.x) |
| Motivated to engage with digital services | XX xx.xx (xx.x) | XX xx.xx (xx.x) |
| Access to digital services that work | XX xx.xx (xx.x) | XX xx.xx (xx.x) |
| Digital services that suit individual needs | XX xx.xx (xx.x) | XX xx.xx (xx.x) |
| **3 Months** |  |  |
| Using technology to process health information | XX xx.xx (xx.x) | XX xx.xx (xx.x) |
| Understanding of health concepts and language | XX xx.xx (xx.x) | XX xx.xx (xx.x) |
| Ability to actively engage with digital services | XX xx.xx (xx.x) | XX xx.xx (xx.x) |
| Feel safe and in control | XX xx.xx (xx.x) | XX xx.xx (xx.x) |
| Motivated to engage with digital services | XX xx.xx (xx.x) | XX xx.xx (xx.x) |
| Access to digital services that work | XX xx.xx (xx.x) | XX xx.xx (xx.x) |
| Digital services that suit individual needs | XX xx.xx (xx.x) | XX xx.xx (xx.x) |
| **Repeat for month 6 and month 12** |  |  |

**Table 5.**
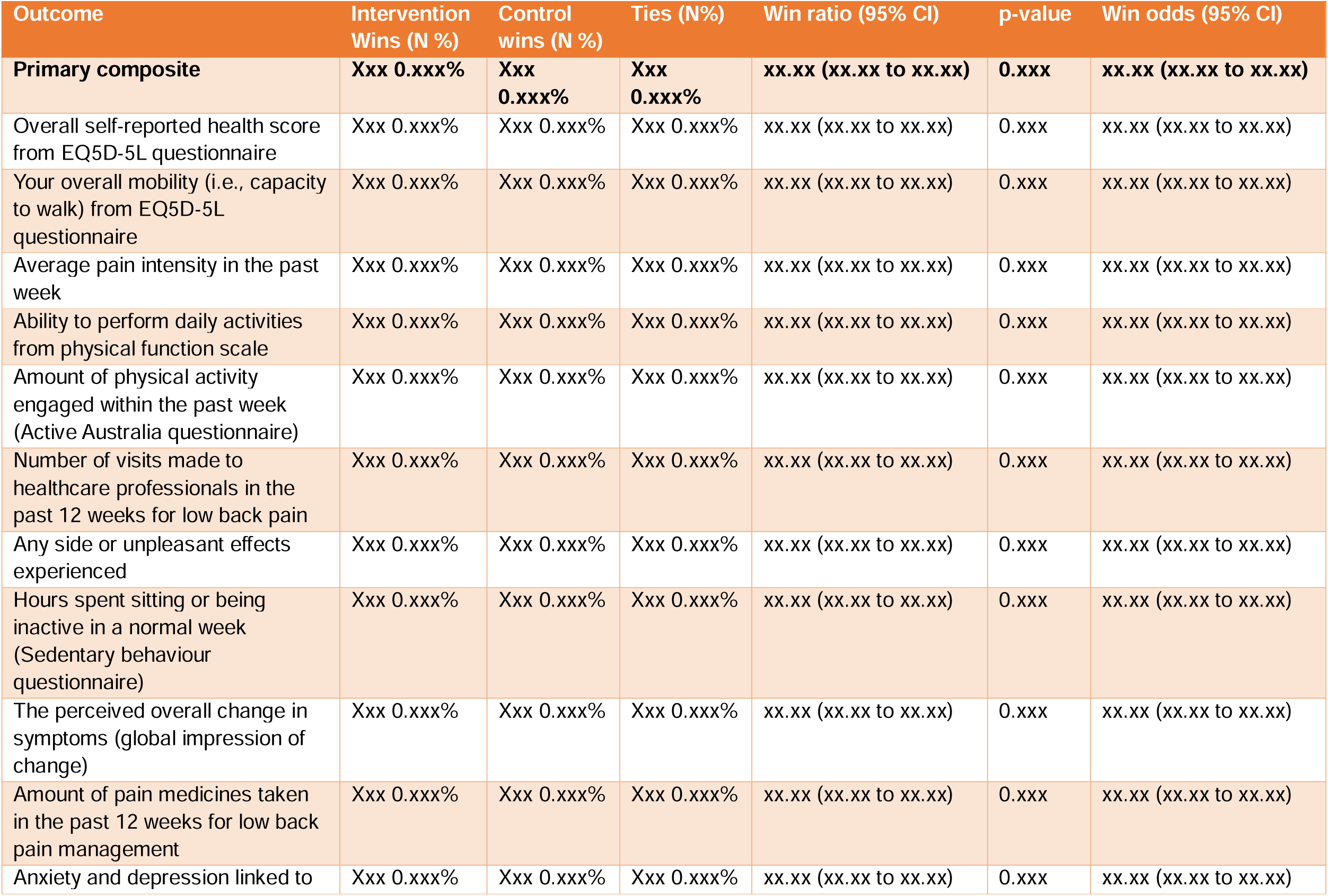
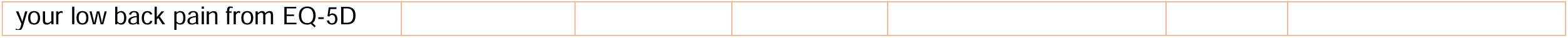
Win ratio analysis of the primary outcome and its components.

**Table 6.** Analysis of secondary outcomes – model results.

| Variable | Intervention<br>Mean (SD) | Control<br>Mean (SD) | MD(95% C.I)<br>/ OR(95%<br>CI)* | p-value |
| --- | --- | --- | --- | --- |
| **Physical function, assessed using the Patient Specific Functional Scale** | Xxx (xx.x%) | Xxx (xx.x%) | xx.xx (xx.x,<br>xx.x) | 0.xxx |
| **Average pain intensity in the past week** | Xxx (xx.x%) | Xxx (xx.x%) | xx.xx (xx.x,<br>xx.x) | 0.xxx |
| **Health utility score from EQ5D** | Xxx (xx.x%) | Xxx (xx.x%) | xx.xx (xx.x,<br>xx.x) | 0.xxx |
| **Hours per week spent in sedentary behaviour** | Xxx (xx.x%) | Xxx (xx.x%) | xx.xx (xx.x,<br>xx.x) | 0.xxx |
| **Participant's global impression of change (ordinal)** | Xxx (xx.x%) | Xxx (xx.x%) | xx.xx (xx.x,<br>xx.x) | 0.xxx* |
| **Physical activity engagement from Active Australia questionnaire (binary)** |  |  |  | 0.xxx* |
| Insufficiently Active (1-149 min/week) | Xxx (xx.x%) | Xxx (xx.x%) | xx.xx (xx.x,<br>xx.x) | 0.xxx |

\*Global p-value

**Table 7.** Exploratory outcomes.

| Variable | Intervention<br>(N = ) | Control<br>(N = ) | Total<br>(N = ) |
| --- | --- | --- | --- |
| **Smallest worthwhile effect achieved at 12 months (yes vs. no)** | Xxx (xx.x%) | Xxx (xx.x%) | Xxx<br>(xx.x%) |
| **Use of hospital, medical and health services for LBP over 12 months** |  |  |  |
| Number of patients | xxx | xxx | xxx |
| Number of events |  |  |  |
| Mean (SD) | xxx.x (xxx.x) | xxx.x (xxx.x) | xxx.x<br>(xxx.x) |
| Median (Q1; Q3) | xxx (xxx; xxx) | xxx (xxx; xxx) | xxx (xxx;<br>xxx) |
| min max | xxx to xxx | xxx to xxx | xxx to<br>xxx |
| Sum | xxx | xxx | xxx |

**Table 8.** Summary of adverse events at all follow-up timepoints.

|  | Intervention<br>(N =xxx ) | Control<br>(N =xxx ) | p-value<br>(1) |
| --- | --- | --- | --- |
| **Adverse drug reactions** | nEVT nPT(xx.x%) | nEVT<br>nPT(xx.x%) | 0.xxx |
| **Serious adverse events (SAE)** | nEVT nPT(xx.x%) | nEVT<br>nPT(xx.x%) | 0.xxx |
| **Serious events** |  |  |  |
| Death or life-threatening | nEVT nPT(xx.x%) | nEVT<br>nPT(xx.x%) |  |
| Causing or prolonging a hospital stay | nEVT nPT(xx.x%) | nEVT<br>nPT(xx.x%) |  |
| Severe disability or incompetence | nEVT nPT(xx.x%) | nEVT<br>nPT(xx.x%) |  |
| Congenital malformations in offspring | nEVT nPT(xx.x%) | nEVT<br>nPT(xx.x%) |  |
| Important clinical events as judged by the investigators | nEVT nPT(xx.x%) | nEVT<br>nPT(xx.x%) |  |
| **SAE severity** |  |  |  |
| Mild (transient, does not interfere with normal activity) | nEVT nPT(xx.x%) | nEVT<br>nPT(xx.x%) |  |
| Moderate (interferes with normal activities) | nEVT nPT(xx.x%) | nEVT<br>nPT(xx.x%) |  |
| Severe (inability to perform daily activities) | nEVT nPT(xx.x%) | nEVT<br>nPT(xx.x%) |  |
| **SAE outcome** |  |  |  |
| Persistent/unresolved | nEVT nPT(xx.x%) | nEVT<br>nPT(xx.x%) |  |
| Resolved but left with sequelae | nEVT nPT(xx.x%) | nEVT<br>nPT(xx.x%) |  |
| Remission | nEVT nPT(xx.x%) | nEVT<br>nPT(xx.x%) |  |
| Fatal | nEVT nPT(xx.x%) | nEVT<br>nPT(xx.x%) |  |
| Unknown | nEVT nPT(xx.x%) | nEVT<br>nPT(xx.x%) |  |

Listing 1. AE listing

**Figure 2: Forest plot for subgroup analysis**

This will be done only for the overall Win ratio analysis. Within each subgroup, it will show the Win Ratio and 95% CI. No p-value will be displayed.

**Figure 3:**
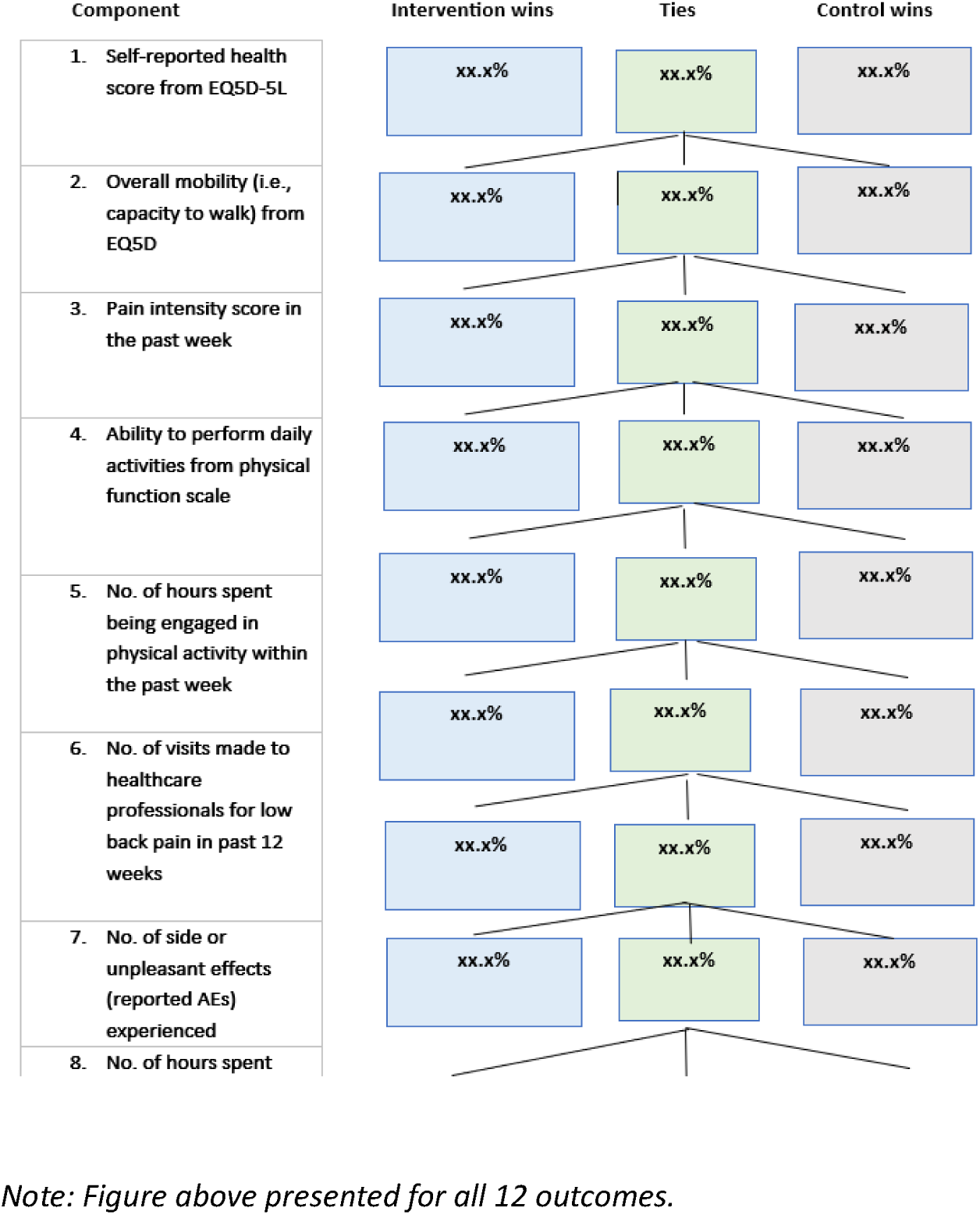
Flow diagram of win ratio decisions.

**Figure 4:**
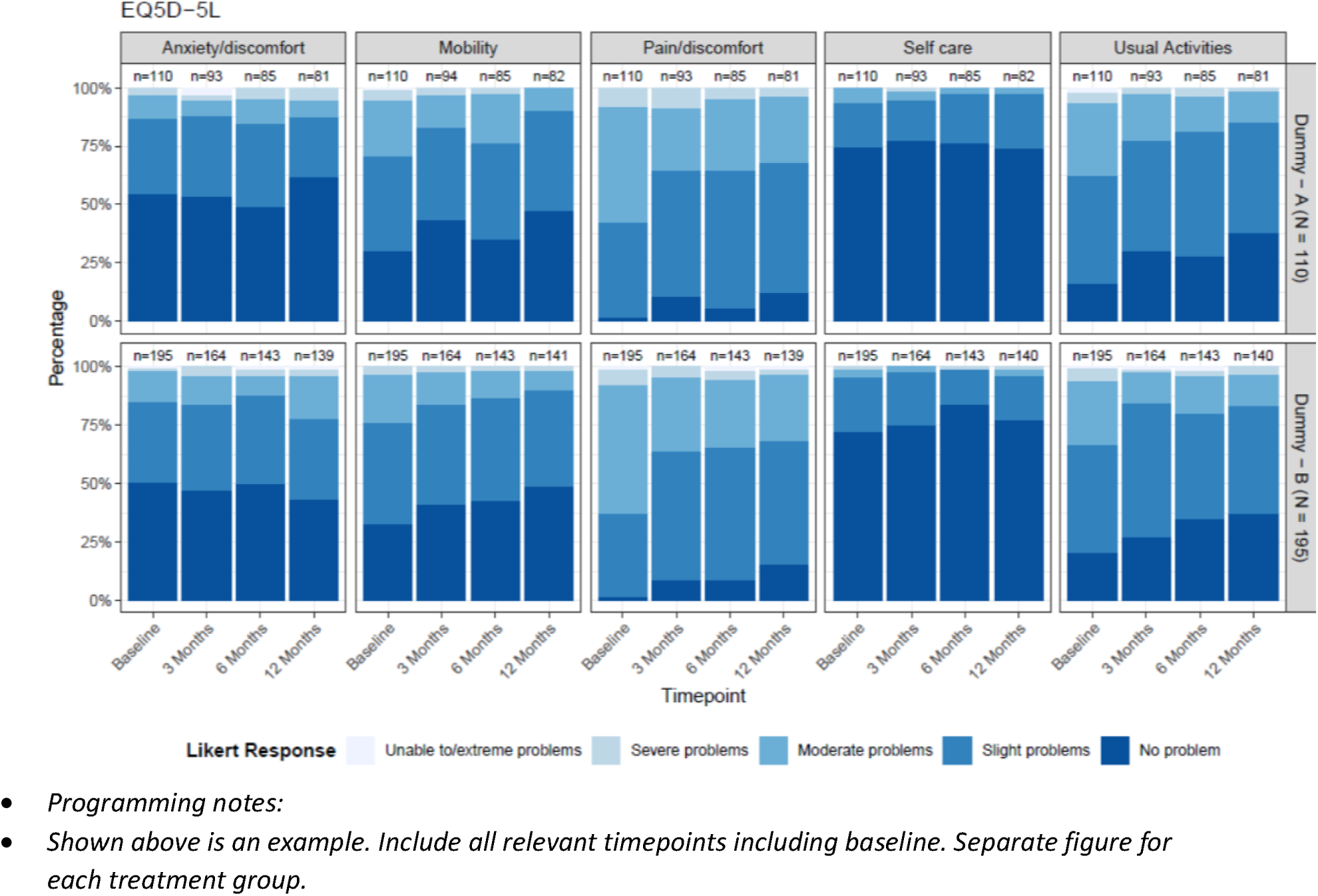
Bar chart of EQ5D-5L across all timepoints (Baseline to 12 months)

**Figure 5: Mean plot of health utility score (EQ5D) over time (Baseline to 12 Months) by treatment groups**

Programming notes:

Display raw means on the graph as numbers near each dot and denominators below the x-axis

**Figure 6: Heatmap of healthcare utilization over time**

